# Repetition-dependent adaptation and Prediction error signalling in Schizophrenia patients with auditory hallucinations: A Roving Mismatch Negativity Study

**DOI:** 10.1101/2023.05.02.23289367

**Authors:** Anushree Bose, Swarna Buddha Nayok, Harsh Pathak, Kiran Basawaraj Bagali, Harleen Chhabra, Satish Suhas, Venkataram Shivakumar, Vanteemar S. Sreeraj, Janardhanan C. Narayanaswamy, Ganesan Venkatasubramanian

## Abstract

**Background:** Prediction error is the surprise that is elicited when the sensory expectations are first established and then violated. Positive symptoms of schizophrenia, like auditory hallucinations (AH), are thought to arise from dysregulated prediction error-signalling. Roving mismatch negativity (rMMN) is a unique event-related potential (ERP) based assessment that allows examination of repetition-dependent adaptation and deviance detection—complimentary processes that are integral to prediction-error signalling. In the rMMN paradigm, the deviant tone becomes the new standard with several repetitions. Also, the number of repetitions of the standard stimuli keeps changing throughout the experiment; longer repetitions yield a more positive ERP response; this phenomenon is Repetition Positivity (RP). Longer repetitions of standard stimuli elicit stronger deviance detection when interrupted, and this is called deviant negativity (DN). A difference waveform between RP and DN reflects the strength of prediction error signalling, the mismatch negativity (MMN).

**Methods:** Twenty-three schizophrenia patients with auditory hallucinations (SZ-AH) and twenty-three healthy controls (HC) underwent rMMN assessment. Standard stimuli were repeated in sets of 3, 8 and 33. The first tone of the succeeding set became the deviant for the preceding set, yielding three components for RP (RP3, RP8, RP33), DN (DN3, DN8, DN33), and MMN (MMN3, MMN8, MMN33). Amplitudes and latencies of these components were compared between SZ-AH and HC. We also looked for potential associations between rMMN indices (amplitudes and latencies) and clinical scores in SZ-AVH. We examined the correlation between the RP-DN pair for all three repetition sets (3, 8, 33).

**Results:** SZ-AH had suppressed DN (DN3, DN8, DN33) and MMN33 amplitudes in comparison to HC. However, none of the RP amplitudes were diminished. Only MMN33 latency was significantly longer in SZ-AH than in HC. Amplitudes and latencies associated with repetition set of 8 showed a significant correlation with the frequency and loudness of AH. HC showed a strong positive correlation between RP-DN pairs; SZ-AH did not, except for the RP33-DN33 pair.

**Discussion:** The link between repetition-dependent sensory adaptation and deviance detection is compromised in SZ-AH patients. Though RP profile (RP3, RP8, RP33) is unimpaired in SZ-AVH, it is potentially due to AH interfering with auditory information processing and not because of intact short-term plasticity of the echoic memory trace.

## INTRODUCTION

Schizophrenia (SZ) is postulated to be associated with aberrant perceptual synthesis underwritten by prediction error signalling deficits. Perceptual synthesis refers to the phenomenon of integrating information and experience from our senses to form knowledge about the external world, inform a safe and profitable engagement with it, and remove redundant or irrelevant information about both (American Psychological Association, 2023). Predictive coding is an active, adaptive phenomenon where based on past (sensory) experience, the brain correctly identifies and screens out unessential information and allows it to focus on unanticipated information to stay up to date with the changes in its sensory environment (Fong et al., 2020). The prediction error is the surprise that is elicited when the sensory expectations are first established and then violated (Fletcher and Frith, 2009). Feedforward models posit that predictive coding is a bi-directional, hierarchical, iterative process of Bayesian inference wherein by integrating the latest sensory information, the brain continually generates and updates its prediction models to prognosticate the environment (Fong et al., 2020). The brain then compares its predictive models to the information/sensations from the incoming stream of stimuli. Neural responses to the incoming stimuli may either match with or violate the apriori prediction model. In case of a match between predicted and incoming signals, the neural response to stimuli received is suppressed, whereas in case of a mismatch, the neural response to the unexpected stimuli elicits a surprise or ‘prediction error’ (Garrido et al., 2009; McCleery et al., 2019). Prediction, thus, informs perception, and prediction error signalling is integral to perceptual synthesis—a faculty noted to be compromised in schizophrenia (Ford and Mathalon, 2012), and especially attributed to the pathogenesis of hallucinations and delusions (Adams et al., 2013; Fletcher and Frith, 2009; Horga et al., 2014).

The predictive coding along the auditory pathway is widely studied with mismatch negativity (MMN). As an electro-encephalography (EEG) derived signature, Mismatch negativity (MMN) is elicited in response to unexpected stimuli when the regularity of a preceding train of oft-repeating stimuli is suddenly interrupted (Näätänen et al., 2007). Mismatch negativity (MMN) is a neurophysiological index of automatic context-dependent change detection that occurs independent of the participant’s conscious attention (Fitzgerald and Todd, 2020). Given the intricate association of SZ pathogenesis with context-depending processing abnormalities (Kirschner et al., 2018) & predictive coding deficits (Friston, 2023) MMN is, therefore, a suitable tool for examining prediction error signalling deficits in Schizophrenia (SZ) patients (Kirihara et al., 2020; Randeniya et al., 2018; Rentzsch et al., 2015; Taylor et al., 2017; Wacongne, 2016).

To capture MMN through a typical auditory event-related potential (ERP) paradigm, at least two components are randomly presented within a passive auditory oddball paradigm: the standard stimuli and the deviant stimuli. The standard stimuli (≥85% presentation) are usually pure tones that are repeated enough times such that they get encoded in short-term echoic (auditory) memory and one comes to expect their occurrence. The deviant stimuli (≤15% presentation) are stimuli that violate the sensory expectation established from the echoic memory trace that the standard stimuli will recur. The deviant stimulus can be a tone that differs from the standard (tone) on one or several acoustic features like pitch/frequency, duration, and intensity/volume (Fisher et al., 2011; Todd et al., 2008). The deviant stimuli can also be a novel sound (like a knock, car horn, etc.) (Fisher et al., 2014) or a gap (unexpected absence of an anticipated tone within a string of tones) (Bertoli et al., 2001; Salisbury and McCathern, 2016) [see (Avissar et al., 2018) for a detailed review on MMN deviant types]. MMN is obtained by subtracting ERP responses to the standard stimuli from the ERP responses to the deviant stimuli. MMN is deemed to be the most negative deflection in scalp-recorded voltage in this difference (deviant—standard) waveform, occurring between 100-240 milliseconds (Umbricht and Krljes, 2005). As the MMN activity is maximal over the frontocentral electrodes, MMN peak amplitude and latency are most frequently reported from the Fz or FCz electrode sites. Despite the variability in deviant types, MMN amplitude is reported to be consistently attenuated in SZ patients in comparison to healthy controls [see metanalyses (Avissar et al., 2018; Erickson et al., 2016)]. MMN abnormalities are noted to precede the onset of schizophrenia (Donaldson et al., 2022; Murphy et al., 2020; Perez et al., 2014) and even predict transition to psychosis from the prodromal state (Hamilton et al., 2022; Näätänen et al., 2015).

However, MMN is not a unitary phenomenon, and two complimentary processes are reported to affect its elicitation: adaptation and deviance detection. Sensory adaptation to the oft-repeating standard stimuli occurs first and sets the expectation for the sensory context. Deviance detection (surprise) follows when this context-dependent sensory expectation is violated (Kirihara et al., 2020) and the prediction model is updated. Predictive coding, as captured by MMN, thus is a nuanced phenomenon that requires concurrent examination of both sensory adaptation and deviance detection in real time in a dynamic sensory context. Roving MMN (rMMN) – a modified version of the MMN task – allows assessment of sensory adaptation and deviance detection; this is achieved by varying the number of repetitions of a standard stimulus, and by using more than one type of standard and deviant stimuli (Baldeweg, 2007; McCleery et al., 2019; McCleery et al., 2018). Within a rMMN paradigm—which is also a passive auditory oddball paradigm—the train of a standard stimulus is interrupted by a deviant stimulus; however, instead of being presented just once, the deviant stimulus is repeated several times such that it becomes the new standard. Furthermore, the length of the standard stimuli sequence varies throughout the experiment such that a longer sequence (that is, greater number of repetitions of) standard stimuli generate a more positive event-related potential (ERP) response; this is called as ‘repetition positivity’ effect. Repetition positivity (RP) is postulated to reflect the stimulus-specific adaptive short-term sensory neuroplasticity (Baldeweg, 2007; McCleery et al., 2018). The RP component of rMMN specifically captures repetition-dependent short-term plasticity in the human brain, and its strength reflects the expectation (prediction) that the standard stimuli will repeat (McCleery et al., 2019). Likewise, the deviant stimulus and resultant MMN that have the longer sequence of preceding standard stimuli, elicit a higher magnitude of deviance detection (Baldeweg et al., 2004; McCleery et al., 2018). Deviance detection increases in magnitude in response to an increase in the length of standard stimuli train preceding it, implying that stronger predictions evoke larger prediction error signals (McCleery et al., 2019). The greater magnitude of MMN reflects the underlying discrepancy between the (adaptive) response to the standard stimuli and (surprise detection) response to the deviant stimuli (Todd and Robinson, 2010).

In addition to examining repetition-dependent short-term plasticity and prediction error signalling, the rMMN paradigm enables probing into the efficiency of echoic memory trace formation (memory trace effect). When the type and repetition length for the standard stimuli is repeatedly changed throughout the experiment, variability is induced in their de novo echoic memory traces (Baldeweg and Hirsch, 2015). These echoic memory traces can be made short (3 repetitions of standard stimuli), intermediate (8 repetitions of standard stimuli), or long (33 repetitions of standard stimuli) (McCleery et al., 2019) and in essence capture short-term auditory plasticity processes (Baldeweg and Hirsch, 2015). For standard stimuli, longer repetitions yield stronger adaptation and expectation that the standard stimuli will recur (more positive ERP component) than shorter repetitions; comparing the long and short RP sub-components reflects the repetition-dependent relative strength of repetition positivity (RP) memory trace effect (RP_MT_). Similarly, for the deviant stimuli, the deviance detection is stronger (more negative ERP component) when a deviant is preceded by a long train of standard stimuli in comparison to a short train of standard stimuli; contrasting the long and short DN sub-components reflects the context-dependent relative strength of deviant negativity (DN) memory trace effect (DN_MT_). And, lastly, the MMN generated by a difference wave of a long repetition train’s deviant and standard pair has higher prediction error signalling (more negative MMN) than an MMN generated from a difference wave of a short repetition train’s deviant and standard pair; this permits us to explore mismatch negativity (MMN) memory trace effect (MMN_MT_) (McCleery et al., 2019).

In this study, we examined repetition-dependent adaptation, deviance detection and prediction error signalling abnormalities in a sample of Schizophrenia patients with significant auditory hallucinations (SZ-AH) using a standard roving MMN paradigm. In line with the existing literature, as RP is reported to be unimpaired in SZ (Coffman et al., 2017; McCleery et al., 2019; McCleery et al., 2018), we did not form any apriori hypothesis about it. We expected SZ-AH patients to have deficient DN and MMN amplitudes in comparison to the healthy control (HC) group. Furthermore, we performed the following exploratory analyses: a) RP, DN and MMN latencies comparison between SZ-AH and HC. b) correlation between RP and DN pairs for each of the repetition sets (3,8,33) in SZ-AH and HC. As RP and DN are interlinked (as a deviant preceded by a higher number of repetitions of standard stimuli elicits a larger deviance detection), we expected a strong correlation between the RP-DN pairs (RP3-DN3, RP8-DN8, RP33-DN33) in HC and but anticipated this relationship to be weak or non-existent in SZ-AH c) whether clinical features (like illness chronicity, medication dose, etc.), psychopathology scores and components of auditory hallucinations (like frequency, reality, loudness, distress, etc.) correlate with amplitudes or latencies of rMMN components of repetition positivity (RP), deviant negativity (DN) or mismatch negativity (MMN) or their memory trace effects (RP_MT_, DN_MT_, MMN_MT_).

## METHODS

### Participants

In this study, twenty-six SZ-AH patients and twenty-three healthy controls were examined. The SZ-AH patients were recruited from the clinical services of the National Institute of Mental Health And Neuro Sciences (NIMHANS), Bengaluru. DSM 5 diagnosis of Schizophrenia was ascertained upon clinical evaluation by senior psychiatrists. All the participants were right-handed and gave written informed consent before participation. This study was approved by the institute ethics committee.

### Clinical assessment

Detailed clinical and treatment history was taken from the patients and caregivers. Scale for assessment of Positive Symptoms (SAPS) (Andreasen, 1984) and Scale for assessment of Negative Symptoms (SANS) (Andreasen, 1989) were used to do a comprehensive assessment of the positive and negative symptom psychopathology in SZ-AH patients. Auditory Hallucination Rating Scale (AHRS) was used for a thorough assessment of hallucinations (Hoffman et al., 2005). AHRS assessment was done before and after add-on acctDCS therapy. The medication dose was noted and has been reported as olanzapine equivalent as per previous guidelines (Leucht et al., 2016). All the participants were screened for conductive and sensorineural hearing loss with Rinne’s and Weber’s test using 512Hz and 256Hz tuning forks before undergoing auditory event-related potential assessment of rMMN.

### Assessment of Roving Mismatch Negativity

#### DATA ACQUISITION

Roving MMN (rMMN) paradigm was administered within an event-related potential experimental protocol as per previous descriptions (McCleery et al., 2019; McCleery et al., 2018) in a sound-attenuated, dimly illuminated, electrically shielded room. The paradigm was designed with E-Prime 3.0 stimulus presentation software (https://pstnet.com/products/e-prime/) and integrated with Brain Vision Recorder EEG acquisition system (https://brainvision.com/products/recorder/) with the help of Chronos stimulus-response device and input-output expander. Auditory stimuli were delivered via BOSE QuietComfort 20 Acoustic headphones (with the noise-cancellation feature off) while subjects watched a nature-themed muted video. The participants were instructed to intently focus on the video and ignore the sounds coming through the earphones. They were asked simple questions related to the video at the end of the experiment to check whether they had paid attention to the video. To ensure clear deliverance of auditory stimuli, the sound intensity was set at 80 dB which was adequately above the auditory threshold of all participants. The rMMN paradigm consisted of eight blocks with a 30 seconds break between two consecutive blocks. The experiment consisted of 1856 stimuli (Stimuli counts: RP3=200, DN3=88, RP8=448, DN8=64, RP33=1024, DN33=32) and took approximately 18 minutes to complete. The stimuli were auditory pure tones of 50ms or 100ms of 700, 800, 900, 1000, 1100, or 1200 Hz. Standard tones were presented as repeating sets of 3 or 8 or 33 in a pseudo-random order within a block. No two consecutive blocks had the same order of stimuli presentation. The deviant tone differed from the preceding tone in both pitch (range: ±100-300) and duration (±50ms), and thus was a double deviant. The deviant tone too was repeated in sets of 3, 8, or 33, and thus became the new standard. Continuous EEG data was recorded at a sampling rate of 1000 Hz from the ActiChamp amplifier without filters. Data was recorded in BrainVision Recorder (Brain Products, GmbH) from 32 active electrodes using EasyCap (EASYCAP GmbH) with pre-marked electrode holders as per the 10-10 electrode placement system. The ground electrode was placed on the forehead. T9 was used as a reference during the acquisition. Ocular activity was recorded with two auxiliary bipolar channels VEOG (vertical electrooculogram) and HEOG (horizontal electrooculogram).

#### DATA PROCESSING

Off-line signal processing was done using BrainVision Analyzer 2.2.2 (Brain Products, GmbH). Data was band pass filtered at 0.5 to 20 Hz. Eye blinks were corrected using Gratton & Coles method (Gratton et al., 1983) using signals from VEOG and HEOG channels. The time segments with movement or other artefacts were excluded from further analysis using a semi-automated (interactive view) artifact rejection algorithm whereby trials were rejected when (a) the absolute voltage difference between two data points exceeded ±100 μV, (b) the difference between the maximum and minimum value in a 200ms segment exceeded ±100 μV, (c) the amplitude exceeded ±100 μV, (d) the difference between the maximum and minimum in an interval of 100 ms exceeded 0.5 μV and (e) deemed noisy upon visual inspection. Artifact-free, continuous EEG data was segmented into 500 msec epochs, from –100 msec to 400 msec relative to the tone onset. The pre-stimulus interval was used for baseline correction. Epochs were separately averaged for all the sets of standard (Repetition Positivity-RP 3, 8 and 33) and deviant tones (Deviant Negativity-DN 3, 8 and 33). It was ascertained that a minimum of 20 artifact-free trials were present for averaging across standards (RP 3, 8 and 33) and deviant (DN 3, 8 and 33) for each participant. MMN waveform was generated by subtracting the standard average (RP) from the corresponding deviant average (DN) separately for sets of 3, 8 and 33. Automated peak detection for the most negative peak was done within a window period of 100 to 200 msec. The peak negative amplitude and its latency were extracted for statistical analyses. Memory trace (MT)—that is, relative strength of the echoic memory trace—was determined separately for RP, DN and MMN by subtracting the amplitude of the shortest train (3^rd^) from the amplitude of the longest train (33^rd^); e.g., MMN33 – MMN3.

At the end of experiment, it was noted whether or not the SZ-AH participants experienced AH during the assessment.

### Statistical Analysis

Statistical analyses were performed in SPSS software (ver. 28). Univariate normality of demographic, clinical and neurophysiological variables was tested using Shapiro-Wilk test (p>0.05) and histograms and outlier detection was done with box plot. Parametric (independent sample t–test, Pearson’s correlation, etc.) or equivalent non–parametric tests (Mann–Whitney U test, spearman’s rank correlation, etc.) were performed to test for statistically relevant groups differences on demographic variables or correlations with clinical scores.

A 2 (Groups: SZ and HC) x 3 (Repetition sets: 3, 8 and 33) MANOVA was performed as a joint test to check for any significant group difference separately for repetition positivity (RP3, RP8, RP33), deviant negativity (DN3, DN8, DN33) and mismatch negativity (MMN3, MMN8, MMN33) amplitudes. Assumptions of MANOVA were tested in the following way: a) multivariate outlier detection was done using Mahalanobis distance (<13.82, df=2), b) multivariate normality was visually tested by plotting matrix scatter plot (∼elliptical distribution was ascertained), c) singularity and multicollinearity within sets of dependent variables (RP-3,8,33; DN-3,8,33; MMN-3,8,33) was ruled out with bivariate Pearson’s correlation (range: r≥0.2 & ≤0.9), d) equality of observed covariance matrices across the groups was ascertained with the Box’s test of equality of covariance of matrices (p>0.05), and e) heteroskedasticity in the dependent variables was ruled out with Levene’s test of equality of error variance (p>0.05). Pillai’s Trace coefficient is considered to be most robust to violations in assumptions when sample sizes are equal (Bray and Maxwell, 1985). We have noted Pillai’s Trace instead of Wilk’s Lambda for the Multivariate tests as a) RP8, DN3 and DN33 for SZ were not normally distributed (Shapiro Wilk p<0.05), and b) MMN3 did not correlate with MMN8 and MMN33. However, all the other above-mentioned assumptions were met. As per previous recommendations (Hsu, 1996)^1^, we checked for significant difference in both multivariate omnibus test and univariate tests even if the omnibus test was not significant. Bonferroni corrected p-value for each of the repetition sets (3, 8, 33) for univariate comparison was set at p=0.016 (that is, 0.05/3).

Additionally, we tested RP, DN and MMN latencies for potential group differences with a 2 (Groups: SZ and HC) x 3 (Repetition sets: 3, 8 and 33) Kruskal Wallis H test. Follow-up Mann Whitney U tests were done to detect which repetition set had significant groups differences. Bonferroni corrected p-value for each of the repetition sets (3, 8, 33) for univariate comparison was set at p=0.016 (that is, 0.05/3).

Given the link between RP and DN, we predicted that in HC the RP-DN pair for all the repetition sets (3, 8 and 33) will show significant correlation with each other, however if RP and/or DN are dysregulated in the SZ-AH group, then RP-DN correlation will be weak or non-existent for them. Bonferroni corrected p-value for RP-DN pair from each of the repetition sets (3, 8, 33) for bivariate correlation was set at p=0.008 (that is, 0.05/6).

Bivariate correlational tests (Pearson’s r and Spearman’s rho) were done to check whether rMMN amplitude or latencies or memory traces (RP_MT_=RP33–RP3; DN_MT_=DN33–DN3; MMN_MT_=MMN33–MMN3) correlated with clinical features like duration of illness, duration of untreated psychosis, clinical scores (Total SAPS, Total SANS, Total AHRS total and AHRS item scores), and medication dose. As the number of these comparisons were too many (21 rMMN indices x 13 clinical indices), instead of a Bonferroni correction, we decided to report only those correlations coefficients that had at least a moderate strength (pearson’s r or spearman’s ρ>0.4), no extreme outlier or no prominent skewness on the scatterplot, and at least a modest best-line fit (R^2^≥0.2) for this segment of exploratory analysis.

## RESULTS

### Baseline Comparison between SZ and HC

Two SZ participants were excluded from the study because of noisy EEG data. For one SZ subject, clinical scores were not available but EEG assessment was completed and demographic details were available. For this patient, group mean was imputed for SAPS, SANS and AHRS. During multivariate analysis, one SZ participant exceeded Mahalanobis distance limit (>13.82, df=2) and was excluded from further analysis.

Demographics details for SZ (n=23) and HC (n=23) groups and clinical details for SZ group are given in Table 1. No significant univariate outliers were detected in any of the neurophysiological, clinical, or demographic variables in SZ and HC groups. The SZ and HC group did not significantly differ from each other on sex distribution (X^**2**^=2.24; p=0.23). However, the SZ participants were significantly less educated than HC (U=414.0; p<0.01) and were older than HC (t=2.38; p=0.02).

**Table 1:** Demographics and clinical variables for SZ-AH and HC.

|  | **Schizophrenia,**<br>**SZ-AH**<br>**(n=23)** | **Healthy**<br>**Controls, HC**<br>**(n=23)** | **Statistics** | **p** |
| --- | --- | --- | --- | --- |
| Age (years) | 33.96±9.54 | 28.70±4.62 | T=2.38 | 0.024* |
| Sex (M:F) | 11:12 | 16:7 | $\chi^2(1)=2.24$ | 0.13 |
| Years of Education<br>(years) | 14.65±1.61 | 16.86±1.98 | U=414 | <0.001* |
| Duration of Untreated<br>Psychosis (months) | 13.91±18.92 | – | – | – |
| Total Duration of Illness<br>(months) | 131.96±112.59 | – | – | – |
| Olanzapine Equivalent | 20.48±12.22 | – | – | – |
| Total SAPS | 30.13±15.45 | – | – | – |
| Total SANS | 30.22±20.21 | – | – | – |
| Total AHRS | 29.43±4.37 | – | – | – |
| AHRS – Frequency | 5.63±2.61 | – | – | – |
| AHRS – Reality | 4.56±0.6 | – | – | – |
| AHRS – Loudness | 2.98±1.01 | – | – | – |
| AHRS – Number of<br>Voices | 3.69±1.94 | – | – | – |
| AHRS – Length | 3.29±1.03 | – | – | – |
| AHRS – Salience | 5.42±1.28 | – | – | – |
| AHRS – Distress | 3.88±1.05 | – | – | – |

\* – Statistical significance at 0.05; $\chi^2$ – Chi-square test; t – T test statistic; U – Mann Whitney U test statistic; p – statistical significance; SAPS=Scale for Assessment of Positive Symptoms; SANS=Scale for Assessment of Negative Symptom; AHRS=Auditory Hallucination Rating Scale

The rMMN amplitude and memory trace for SZ (n=23) and HC (n=23) are given in table 2, and rMMN latencies are given in Table 3. The 2×3 MANOVA of rMMN amplitudes revealed that group type (SZ vs. HC) did not have any significant effect on combined Repetition Positivity (RP3, RP8, or RP33) variables (Pillai’s V=0.004; F_3,42_=0.54; p=0.98; partial η^2^=0.004; observed power=0.059). Univariate tests were not significant for any of the RP variables. SZ patients significantly differed from HC (Pillai’s V=0.19; F_3,42_=3.31; p=0.03; partial η^2^=0.19; observed power=0.71) on combined Deviant negativity variables (DN3, DN8, DN33). Univariate testing found the effect to be significant for DN3 (F=7.42; p<0.01; partial η^2^=0.14), DN8 (F=7.28; p=0.01; partial η^2^=0.14) and DN33 (F=6.07; p=0.018; partial η^2^=0.12). With respect to the combined MMN variables (MMN3, MMN8, MMN33), a significant effect of group type was noted (Pillai’s V=0.18; F_3,42_=3.10; p=0.04; partial η^2^=0.18, observed power=0.68). Univariate test found the effect to be significant only for MMN33 (F=7.97; p<0.01; partial η^2^=0.15).

**Table 2:** Comparison of Roving MMN amplitudes and latencies for SZ-AH and HC.

|  | **SZ-AH (n=23)**<br>**Mean±SD** | **HC (n=23)**<br>**Mean±SD** | **Statistics** | **p** | **$\eta^2$** |
| --- | --- | --- | --- | --- | --- |
| RP3 Amplitude (μV) | -1.97±1.26 | -2.04±1.89 | $F_{3,42}=0.54$ | 0.98 | 0.004 |
| RP8 Amplitude (μV) | -1.38±1.51 | -1.44±1.54 |  |  |  |
| RP33 Amplitude (μV) | -0.9±1.26 | -0.86±1.29 |  |  |  |
| DN3 Amplitude (μV) | -1.87±2 | -3.45±1.94 | $F_{3,42}=3.31$ | 0.03* | 0.19 |
| DN8 Amplitude (μV) | -2.53±1.86 | -3.93±1.65 |  |  |  |
| DN33 Amplitude (μV) | -3.37±2.91 | -5.36±2.54 |  |  |  |
| MMN3 Amplitude (μV) | -1.53±2.08 | -2.44±1.78 | $F_{3,42}=3.10$ | 0.04* | 0.18 |
| MMN8 Amplitude (μV) | -2.62±1.6 | -3.23±1.34 |  |  |  |
| MMN33 Amplitude (μV) | -3.46±2.07 | -5.38±2.52 |  |  |  |
| RP<sub>MT</sub> | 1.07±1.01 | 1.18±1.79 | $t=-0.245$ | 0.81 | - |
| DN<sub>MT</sub> | -1.5±3.19 | -1.91±1.99 | $t=0.52$ | 0.61 | - |
| MMN<sub>MT</sub> | -1.94±2.54 | -2.94±3.16 | $t=1.19$ | 0.24 | - |

\*=Significant at $p=0.05$ ; F=Fisher's Test (MANOVA); p=significance level; $\eta^2$ =eta square; t=independent sample t-test; RP=Repetition Positivity; DN=Deviant Negativity; MMN=Mismatch Negativity; RP<sub>MT</sub>=RP Memory Trace Effect; DN<sub>MT</sub>=DN Memory Trace Effect; MMN<sub>MT</sub>=MMN Memory Trace Effect.

**Table 3:** Comparison of Roving MMN latencies for SZ-AH and HC.

|  | Schizophrenia, SZ-AH (n=23) |  | Healthy Controls, HC (n=23) |  | Statistic | p |
| --- | --- | --- | --- | --- | --- | --- |
|  | Mean±SD | Mean Rank | Mean±SD | Mean Rank |  |  |
| RP3 Latency (msec) | 134.83±37.53 | 21.63 | 141.43±33.82 | 25.37 | $\chi^2(1)=2.51$ | 0.11 |
| RP8 Latency (msec) | 133.7±35.21 | 20.07 | 153.65±37.55 | 26.93 |  |  |
| RP33 Latency (msec) | 133.7±37.24 | 20.39 | 151.48±38.45 | 26.61 |  |  |
| DN3 Latency (msec) | 132.57±35.21 | 22.02 | 134.04±27.06 | 24.98 | $\chi^2(1)=2.97$ | 0.08 |
| DN8 Latency (msec) | 143.91±34.51 | 24.52 | 138.43±27.38 | 22.48 |  |  |
| DN33 Latency (msec) | 134.52±26.59 | 20.09 | 148.48±26.86 | 26.91 |  |  |
| MMN3 Latency (msec) | 145.65±33 | 24.26 | 140.74±29.56 | 22.74 | $\chi^2(1)=6.33$ | 0.01* |
| MMN8 Latency (msec) | 146.43±36.11 | 24.07 | 139.48±23.48 | 22.93 |  |  |
| MMN33 Latency (msec) | 134.26±30.24 | 18.52 | 154.09±21.22 | 28.48 |  |  |

\*=Significant at p=0.05; $\chi^2$ =Kruskal Wallis H Test; p=significance level; RP=Repetition Positivity; DN=Deviant Negativity; MMN=Mismatch Negativity

The Repetition Positivity (RP) profile was comparable between SZ and HC. The expected RP profile is supposed to be RP_3_< RP_8_< RP_33_ in the order of increasing positivity of N1 amplitude; the greater the number of repetitions, the stronger the memory trace and the less negative/more positive the N1 amplitude. The reverse is true for Deviant Negativity (DN) profile and Mismatch Negativity (MMN) profile; the expected pattern is DN_3_< DN_8_<DN_33_ and MMN_3_<MMN_8_<MMN_33_ in the order of increasing negativity of N1 amplitude. The magnitude of prediction error signalling is higher when the train of preceding standard stimuli is longer. Though this pattern is not violated in SZ, in comparison to HC, SZ-AH have prominent attenuation of amplitudes for the entire Deviant Negativity profile (DN3, DN8, DN33) and selective attenuation of amplitudes in the Mismatch Negativity profile, especially MMN33 (Figure 1).

**Figure 1:**
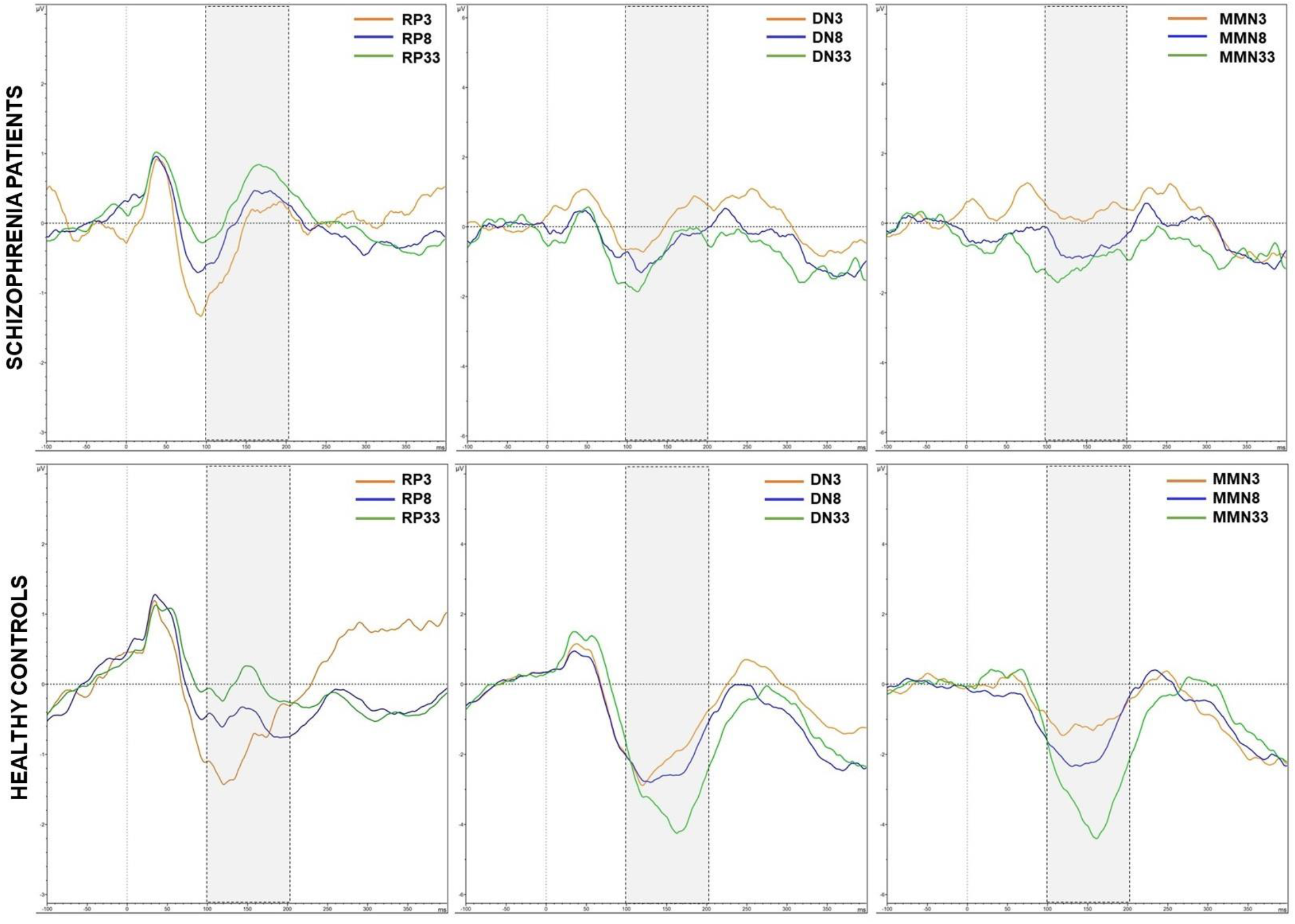
Grand Average waveform for Repetition Positivity (RP3, RP8, RP33), Deviant Negativity (DN3, DN8, DN33), and Mismatch Negativity (MMN3, MMN8, MMN33) profiles for SZ-AH (n=23) and HC (n=23). Figures 1(a) and 1(d) show Repetition Positivity profiles in SZ-AH and HC respectively (amplitude scale: +3 to -3 μV). Figures 1(b) and 1(e) show Deviant Negativity profiles in SZ-AH and HC respectively (amplitude scale: +5 to -6 μV). Figures 1(c) and 1(f) show Mismatch Negativity profiles in SZ-AH and HC respectively (amplitude scale: +5 to -6 μV). SZ-AH have prominent attenuation of amplitudes for the entire Deviant Negativity profile (DN3, DN8, DN33) in comparison to the healthy controls. SZ-AH also show selective attenuation of amplitudes in the Mismatch Negativity profile, especially MMN33.

We performed a 2 (Groups: SZ and HC) x 3 (Repetition sets: 3, 8 and 33) MANOVA with age as covariate (we did not use education years as a covariate since the performance in rMMN experiment is not dependent on formal education skills). With MANCOVA, no effect of group type was noted for combined Repetition Positivity (RP3, RP8, or RP33) variables (Pillai’s V=0.002; F_3,41_=0.027; p=0.99; partial η^2^=0.002) after controlling for age. The univariate tests were also not significant for any of the RP variables. After controlling for age, the group type had no significant effect on combined Deviant negativity (DN3, DN8, DN33) variables (Pillai’s V=0.16; F_3,41_=2.62, p=0.06, partial η^2^=0.16). However, the univariate pair-wise tests were significant for DN3 (F=5.97; p=0.02; partial η^2^=0.12), DN8 (F=5.35; p=0.03; partial η^2^=0.11) and DN33 (F=5.05; p=0.03; partial η^2^=0.11). Likewise, no significant effect of group type was noted for combined MMN (MMN3, MMN8, MMN33) variables (Pillai’s V=0.16; F_3,41_=2.52, p=0.07, partial η^2^=0.16), however, univariate pair-wise test for MMN33 was noted to be significant (F=6.68; p=0.01; partial η^2^=0.13). As a covariate, age had no significant effect on the adjustment of dependent variables of Repetition Positivity (Pillai’s V=0.008; F_3,41_=0.113; p=0.95; partial η^2^=0.008), Deviant Negativity (Pillai’s V=0.010; F_3,41_=0.145; p=0.93; partial η^2^=0.010), and Mismatch Negativity (Pillai’s V=0.030; F_3,41_=0.43; p=0.73; partial η^2^=0.030) in multivariate general linear model.

Memory trace effects for RP, DN and MMN were compared between the two groups. No significant difference was noted for RP_MT_, DN_MT_ and MMN_MT_ in SZ-AH compared to HC. However, though not significant, MMN_MT_ for HC was notably higher than SZ-AH (Table 2).

The 2×3 Kruskal Wallis H test revealed that group type (SZ vs. HC) did not have any significant effect on latencies of combined Repetition Positivity [X^**2**^(1)=2.51; p=0.11] and combined Deviant Negativity [X^**2**^(1)=2.97; p=0.08] (Table 2). However, Kruskal-Wallis H test showed that there was a statistically significant difference in combined Mismatch negativity latencies between the SZ and HC groups [X^**2**^(1)=6.33; p=0.01]. A follow-up Mann Whitney U test revealed MMN33 latency significantly differed between SZ and HC (U=379; p=0.012).

As anticipated, in SZ-AH group the RP-DN correlation was disrupted. They did not show significant correlation between RP and DN components for repetition set of 3 and 8, and had significant positive correlation only between RP33 and DN33. For the HC group, strong positive correlation between RP-DN pair was noted for all the repetition sets (3, 8, 33) (Table 4).

**Table 4:** Correlation between Repetition Positivity and Deviant negativity pairs for repetition sets of 3, 8 and 33.

|  | **SZ-AH (n=23)** | **HC (n=23)** |
| --- | --- | --- |
| **RP3-DN3** | Spearman's $\rho=0.34$<br>$p=0.11$ | Pearson's $r=0.65^*$<br>$p=0.001$ |
| **RP8-DN8** | Spearman's $\rho=0.46$<br>$p=0.02$ | Pearson's $r=0.65^*$<br>$p=0.001$ |
| **RP33-DN33** | Spearman's $\rho=0.63^*$<br>$p=0.001$ | Pearson's $r=0.56^*$<br>$p=0.006$ |

\*=significant at Bonferroni corrected $p \leq 0.008$ ; RP=Repetition Positivity; DN=Deviant Negativity; $p$ =significance level

Frequency of auditory hallucination (AHRS item no. 1) showed significant positive correlation with RP3 amplitude (r=0.59; p<0.01), RP33 (r0.50=; p=0.02) amplitude and DN8 (r=0.45; p=0.03) amplitude such that higher frequency of auditory hallucination co-occurred with more positive peaks of repetition positivity and deviant negativity [Figure 2(a), 2(b) and 2(c)]. Also, frequency of auditory hallucination showed significant negative correlation with MMN33 latency (ρ=-0.57; p<0.01), that is, higher frequency of auditory hallucinations occurred with lower Mismatch Negativity latency [Figure 2(d)]. Loudness of Voices (AHRS item no. 3) showed significant negative correlation with DN8 peak amplitude (ρ=-0.56; p<0.01); louder the auditory hallucinations, more negative the deviant negativity 8 peak amplitude latency [Figure 2(e)].

**Figure 2:**
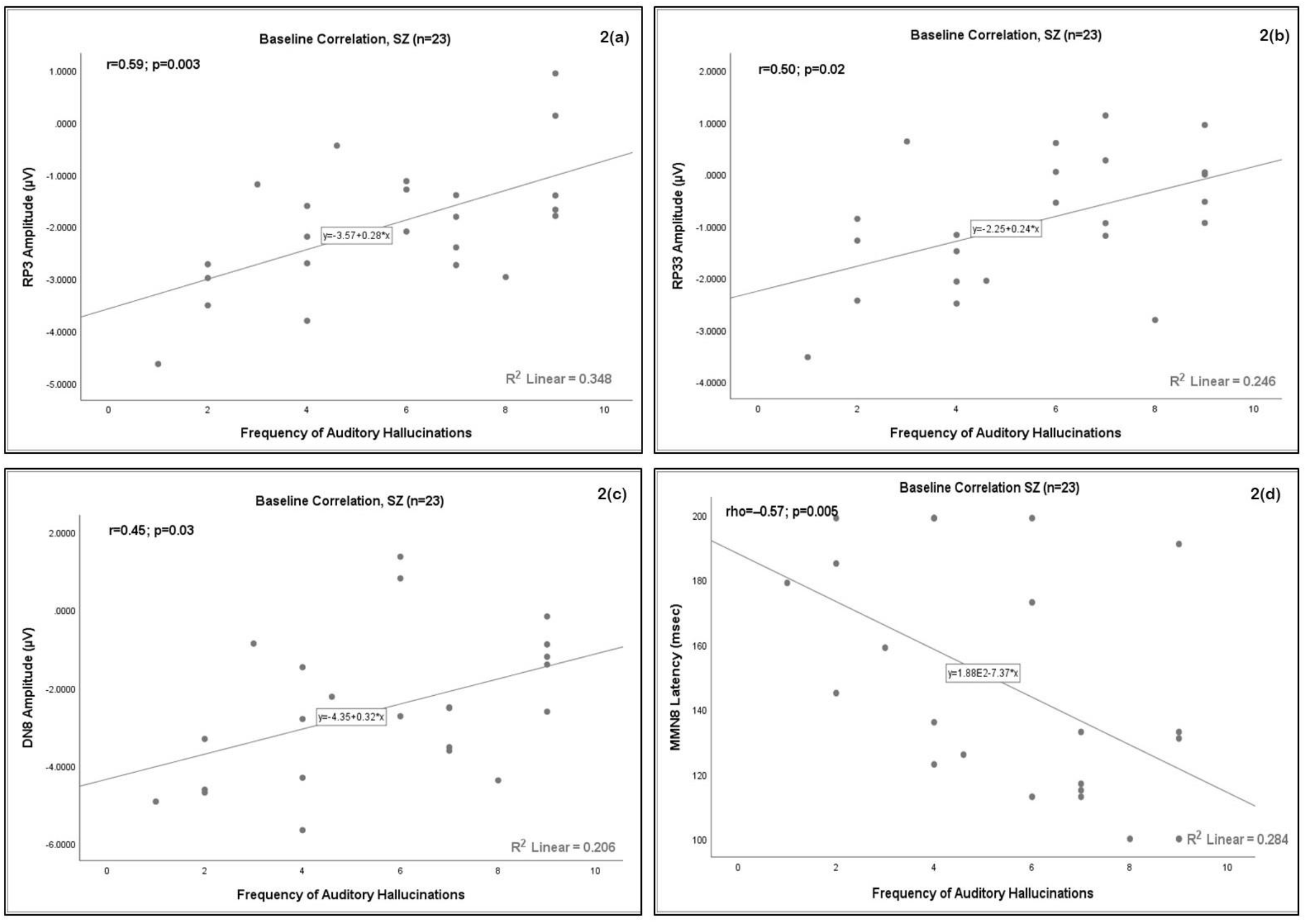
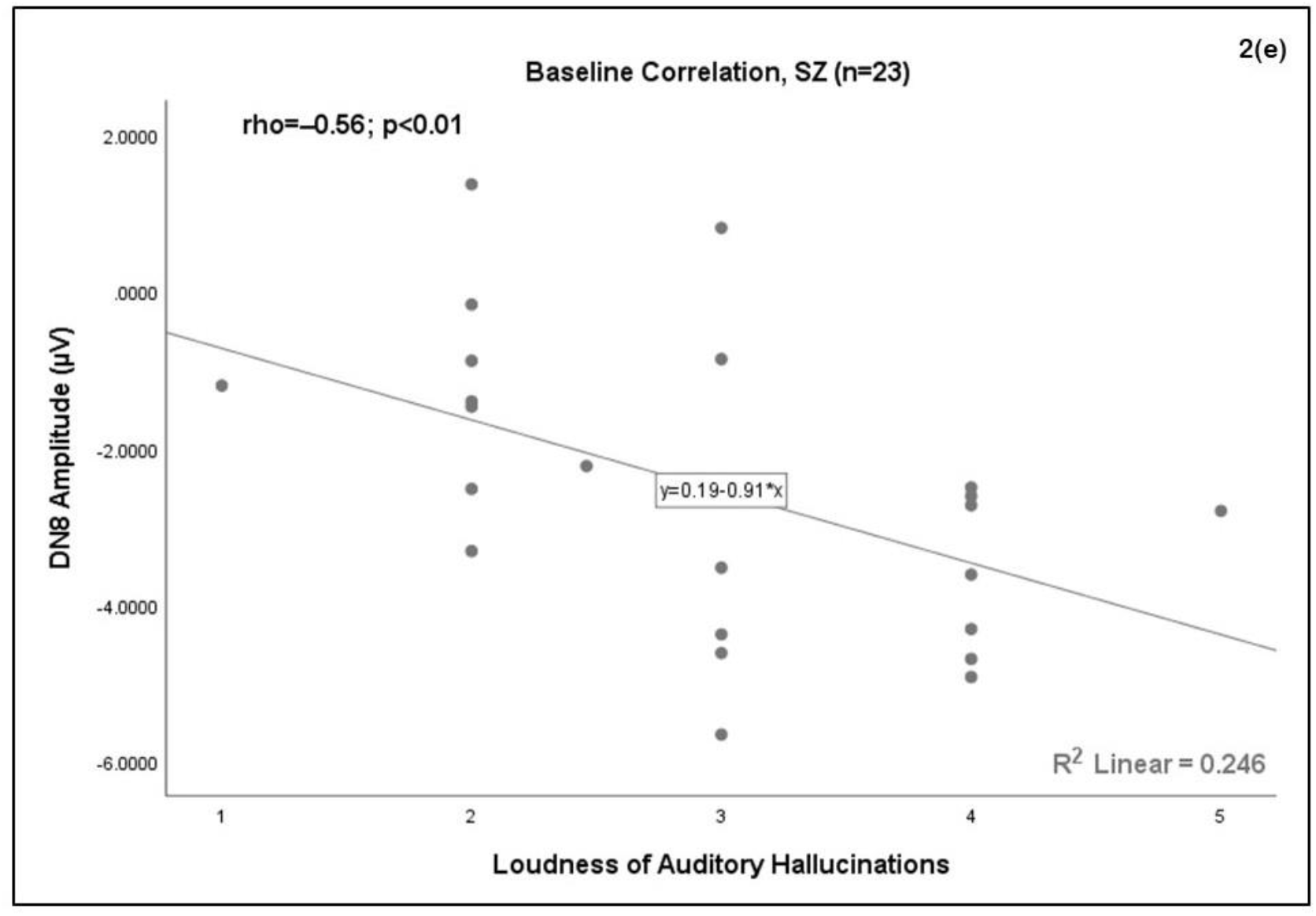
Frequency of auditory hallucinations positively correlated with amplitude of 2(a) Repetition Positivity 3 peak amplitude, 2(b) Repetition Positivity 33 peak amplitude and 2(c) Deviant Negativity 8 peak amplitude; higher frequency of auditory hallucination co-occurred with more positive peaks of repetition positivity and deviant negativity. Frequency of auditory hallucinations negatively correlated with 2(d) Mismatch Negativity 8 Latency; higher frequency of auditory hallucinations occurred with lower Mismatch Negativity latency. Loudness of auditory hallucination negativity correlated with Deviant Negativity 8 amplitude; louder the auditory hallucinations, more negative the deviant negativity peak.

## DISCUSSION

In this study, we aimed to examine repetition-dependent adaptation and prediction error signalling abnormalities in SZ-AH patients and HC using a standard roving MMN paradigm. All patients had experienced AH at some point during the ERP assessment. As anticipated, the SZ-AH patients did not differ from HC on any measure of Repetition Positivity (RP3, RP8, RP33) in concurrence with previous studies (Coffman et al., 2017; McCleery et al., 2019; McCleery et al., 2018). The strongest support, however, comes from McCleery et al., (2019, 2018) because of the similarity in rMMN paradigm used for assessment in their and our studies. One study reported repetition-related adaptation (repetition suppression) to be significantly impaired in SZ compared to HC (Rentzsch et al., 2015); however this study had separately assessed MMN and repetition suppression using two different paradigm unlike us. We used a single test to derive indices for both prediction error signalling and repetition-dependent adaptation rMMN. Comparable RP profile between SZ-AH and HC indicates that in SZ-AH patients the ability to form repetition dependent memory traces is not impaired. Both SZ-AH patients and HC demonstrated the expected RP amplitude profile of RP3<RP8<RP33 in the order of increasing positivity, that is, RP33 is the most positive amplitude among the three because it has strongest echoic memory trace from longest repetition (33) train of the standard stimuli, followed by RP8 and RP3 respectively.

As hypothesized, SZ-AH patients had significantly attenuated amplitude in comparison to HC for all the repetition sets, that is, DN3, DN8 and DN33 which persisted after controlling for age in the multivariate general linear model. However, SZ-AH patients had significantly attenuated MMN only for the repetition set 33 (MMN33); MMN3 and MMN8 amplitudes were comparable between SZ-AH and HC groups. It is interesting to note that SZ-AH showed the correct DN and MMN amplitude profile—which are, DN3<DN8<DN33 & MMN3<MMN8<MMN33 in the order of increasing negativity—just like HC, however, amplitude for DN3, DN8, DN33 and MMN33 components was significantly lower for the SZ-AH in comparison to HC. This finding is different from previous reports using rMMN paradigm with SZ-AH and SZ patients (McCleery et al., 2018) where SZ-AH patients did not show the DN3<DN8<DN33 pattern of increasingly negative amplitude unlike HC and SZ groups. This could be due to differences in sample size and demographics between this study and the present study. Our SZ-AH (n=23, Age: M=33.96, SD=9.54 years) and HC (n=23, Age: M=28.70, SD=4.62 years) groups were larger in size and younger than SZ-AH (n=16, Age: M=51.13, SD=10.23) and HC (n=18, Age: M=49.58, SD=8.99) groups reported by McCleery et al., 2018. Also, differences in ethnicity of participants across these studies may have played a role. Our sample was homogenous (all South-Asian) but theirs’s (17-Caucasian, 12-African-American, 5-Asian, 2-Others) was not.

In line with previous reports (McCleery et al., 2019; McCleery et al., 2018), we did not find memory trace effects for RP, DN or MMN to significantly differ between the groups though MMN_MT_for HC was higher than SZ-AH. For our sample, this could be because the RP components and MMN components, except for MMN33, did not significantly differ between the groups. Furthermore, though DN components significantly differed between the group, the DN amplitude profile of DN3<DN8<DN33 of increasingly negative amplitudes was intact in SZ-AH. Also, the RP and MMN amplitude profiles were not violated. Thus, it is not surprising that the memory trace effects were unimpaired in this study’s SZ-AH sample.

We compared latencies for all of the rMMN components though previous studies have not done so (McCleery et al., 2019; McCleery et al., 2018) probably because latency findings are less straightforward to interpret. Only MMN33 latency was found to be significantly lower for the SZ-AH group. In conjunction with diminished MMN33 amplitude, the conclusion is MMN33 has significantly smaller amplitude and shorter latency in SZ-AH compared to HC. The shorter processing time (latency), may have contributed to inadequate prediction error estimate (smaller amplitude).

RP and DN amplitudes for each of the repetitions sets (3,8, 33) are tightly interlinked. As the magnitude of deviance detection is contingent upon the strength of repetition-dependent sensory adaptation, we expect a tight (↓ scatter) and strong correlation between RP3-DN3, RP8-DN8 and RP33-DN33 pairs. This was true for HC but not for SZ-AH who showed strong correlation only for one RP-DN pair: RP33-DN33. Therefore, it may be concluded that the complementary mechanisms of repetition-dependent adaptation and prediction-error signalling are not working in tandem in SZ-AH patients.

We did not find any significant association between RP, DN or MMN components and gross (total) clinical scores (positive symptoms, negative symptoms, or auditory hallucination), medication dose or illness chronicity after applying our selection criteria [moderate strength of correlation coefficient (r/ρ>0.4), no extreme outlier, no prominent skewness on the scatterplot, and a modest best-line fit (R^2^≥0.2)]. This finding is similar to previous reports where weak or low correlations were noted between clinical score and amplitudes of rMMN components that did not survive multiple corrections (McCleery et al., 2019; McCleery et al., 2018). In fact, even in broader MMN literature, there’s lack of support for correlation between duration deviant and frequency deviant MMN and clinical symptoms, illness chronicity [see metanalyses (Erickson et al., 2017; Erickson et al., 2016)]. A few positive reports do exist for correlation between negative symptoms of SZ and MMN amplitude (Kim et al., 2020; Murphy et al., 2020) and correlation between auditory hallucinations and MMN amplitude (Fisher et al., 2011; Fisher et al., 2012), there’s considerable variability in type of MMN paradigm, clinical scales used for scoring, and SZ sample characteristics. Additionally, findings from neuroimaging study indicate how MMN dysregulation in SZ progressive but non-linear (Salisbury et al., 2007); as a result attenuated MMN amplitude is notable in SZ population independent of illness chronicity and severity of symptoms (Erickson et al., 2016). This explains the inconsistent finding of correlation between MMN indices and psychopathology scores in SZ population.

Perhaps the most intriguing finding of this study is the positive correlation between RP3, RP33 and DN8 amplitudes and frequency of auditory hallucination such that increase in frequency of hallucination associated with more positive RP3, RP33 and DN8 amplitudes. In SZ-AH patients, the RP profile was comparable to HC and it exhibited the expected pattern of increasing positivity of amplitude (RP3<RP8<RP33) with increase in the number of repetitions for standard stimuli. This profile captures the brain’s ability to discount irrelevant sensory information and reflects repetition-dependent, context-specific sensory adaptation. However, it is possible that SZ-AH patients are hyper-attuned to internal percepts (like AH) as opposed to external percepts (auditory tones), and this bias possibly increases with an increase in the frequency of AH. Therefore, it could be that the uncompromised RP profile in SZ-AH is due to a subpersonal (potentially unintentional) preferential bias in information processing rather than due to an intact repetition dependent sensory adaptation. Additionally, DN8 amplitude also showed a positive correlation with frequency of auditory hallucination such that increase in frequency of hallucination associated with more positive DN8 amplitude. SZ-AH showed significantly attenuated DN8 amplitude in comparison to HC. It seems that with an increase in frequency of AH and associated information processing bias towards internal percepts, SZ-AH may have increasingly supressed deviance detection response with respect to DN8. Taken together, this hypothesis of information processing bias leading to unaffected RP amplitudes but diminished DN amplitudes also explains the lack of overall association between RP-DN pairs in SZ-AH.

MMN8 latency showed negative correlation with frequency of auditory hallucination such that lower in frequency of hallucination associated with longer MMN8 latency, that is, longer information processing time. As MMN8 is the difference waveform derived by subtracting ERP response to RP8 from ERP response to DN8, the MMN8 latency reflects the information processing time taken to detect deviance (DN8) relative to the train of repetition of standard stimuli (RP8) preceding it. This implies that SZ-AH with a lower frequency of AH took more time to process prediction error signalling (MMN8) for the repetition set 8 (combined ERP responses to RP8-DN8). Also, SZ-AH with higher frequency of AH gave less time to processing prediction error signal to repetition set 8. The overall group mean for MMN8 latency is higher for SZ-AH than HC, though not significantly. Furthermore, as both MMN8 amplitude and MMN8 latency is not significantly different between SZ-AH and HC, it is likely that SZ-AH needed longer processing time (MMN8 latency) to elicit an adequate prediction error signal (MMN8 amplitude). However, correlation between shorter MMN8 latency and higher frequency of AH indicates that higher AH psychopathology interferes with and compromises auditory information processing; an observation partially corroborated by an overall suppressed DN8 amplitude in SZ-AH and association between suppression of DN8 and frequency of AH. Altogether, this finding lends further support to the above-mentioned theory that correlation between frequency of AH and impairment in deviance detection (DN8) is likely due to biased auditory information processing, and though RP profile in SZ-AH is noted to be not dysregulated, it is probably so not because of adequate repetition dependent sensory adaptation but likely due to a suboptimal auditory information processing stemming from potential preferential bias towards processing internal percepts.

DN8 amplitude also showed negative correlation with loudness of auditory hallucination, such that louder auditory hallucinations associated with diminished (less negative) DN8 amplitudes. It is possible that louder AH stress-out the auditory information processing system such that its responsivity is suppressed or compromised. It is interesting that higher frequency of and louder AH supressed deviance detection for intermediate repetition train (DN8). Also, the prediction error signalling (MMN8) latency for the intermediate repetition was found to shorter indicating inadequate time was given to process deviance detection relative to the repetition-dependent suppression of auditory processing response. Several correlations between features of AH (frequency, loudness) and components of intermediate repetition set (DN8, MMN8), could potentially indicate that the intermediate repetition train is more sensitive to psychopathology than short (3) or long (33) repetition trains. It is hard to speculate why similar association was not observed for repetition train 33, where the magnitude of RP and DN are higher; maybe the results hint towards the possibility of a ‘Goldilock’s zone’ that begs scientific curiosity and replication in a larger sample.

There are several limitations of this study. The overall results were not corrected for multiple comparisons though we have taken care to report only robust findings for exploratory correlation analyses. MMN indices often correlate with measures functional outcomes (Lee et al., 2014; Wynn et al., 2010) and cognitive functions (Baldeweg and Hirsch, 2015) in SZ. As we did not assess these domains, this is a lost opportunity. We did not have a follow-up component as it was beyond the scope of this study. However, given the variability in reports of correlation between clinical symptoms and MMN indices, it would have been interesting to see how correlation between clinical symptoms and MMN indices change over the illness course. High density EEG recordings, if done, could answer whether the cortical generators of RP, DN and MMN vary across SZ-AH and HC. Having additional control groups of SZ patients without hallucination, unaffected SZ siblings, schizotypy patients and unmedicated SZ patients could hightlight similarity and differences in the composite roving MMN profiles (RP, DN, MMN) and indicate whether these deficits are expressed selectively or in a continuum, and whether these are trait or state dependent deficits in schizophrenia spectrum disorders.

To the best of our knowledge, this is the first report to examine the link between repetition-dependent sensory adaptation (RP) and deviance detection (DN) in a graded manner and show this crucial link to be compromised in SZ-AH. Furthermore, though previous studies have reported Repetition Positivity to be preserved in SZ and SZ-AH, with the help of correlation between several RP, DN and MMN indices and features of auditory hallucinations, we are the first to report that the Repetition Positivity profile, though intact, is dysregulated in SZ-AH. We also note that rMMN indices from intermediate repetition set (of 8) are more sensitive to psychopathology scores than short or long repletion sets. However, future studies with larger samples should replicate these findings before drawing conclusions. Additionally, we re-affirm previous findings of compromised deviance detection and prediction-error signalling in SZ-AH. In summary our findings add further evidence to the growing literature on the predictive processing model for schizophrenia (Adams et al., 2013; Sterzer et al., 2018)

## Data Availability

Data from this study are not publicly available due to ethical and privacy restrictions. However, data will be made available upon reasonable request from the corresponding authors.

## CREDIT AUTHOR STATEMENT

Author GVS and AB designed the study. Authors AB, SS, SBN, HP, and KB collected the clinical data. Authors GV & VSS supervised the clinical assessment. ERP experiment was designed and implemented by AB and HC. ERP data was acquired by AB and SBN. AB and SBN did the data entry. AB analysed the ERP data and performed the statistical analyses. Authors AB & GVS managed the literature search and wrote the first draft of manuscript. All authors revised and optimized further versions of the manuscript. All the authors have contributed to and have approved the final manuscript.

## ACKNOWLEDGEMENTS

Anushree Bose is supported by Department of Biotechnology (DBT)-Wellcome Trust India Alliance Early Career Fellowship grant (IA/CPHE/19/1/504591). Swarna Buddha Nayok acknowledges the support of the Indian Council of Medical Research (ICMR). Harsh Pathak and Kiran Basawaraj Bagali are supported by the Department of Biotechnology (DBT) - Wellcome Trust India Alliance (IA/CRC/19/1/610005). Venkataram Shivakumar acknowledges the support of Department of Biotechnology (DBT) - Wellcome Trust India Alliance Early Career Fellowship grant (IA/CPHE/18/1/503956). Vanteemar S Sreeraj acknowledges the support of the India-Korea joint program cooperation of science and technology by the National Research Foundation (NRF) Korea (2020K1A3A1A68093469), the Ministry of Science and ICT (MSIT) Korea, and the Department of Biotechnology (India) (DBT/IC-12031(22)-ICD-DBT). Ganesan Venkatasubramanian acknowledges the support of Department of Biotechnology (DBT) - Wellcome Trust India Alliance (IA/CRC/19/1/610005) and Department of Biotechnology, Government of India (BT/HRD-NBA-NWB/38/2019-20(6)).

## Footnote

1 – “*An unfortunate common practice is to pursue multiple comparisons only when the null hypothesis of homogeneity is rejected*.” (Hsu, page 177). Multiple comparisons can find significant differences between group means even if the overall MANOVA has a p>0.05. With one exception (the test of protected Fisher LDS) such multiple comparisons tests can be valid.

